# PERFECT: Personalized Exercise Recommendation Framework and architECTure

**DOI:** 10.1101/2023.09.14.23295561

**Authors:** Milad Asgari Mehrabadi, Elahe Khatibi, Tamara Jimah, Sina Labbaf, Holly Borg, Pamela Pimentel, Nikil Dutt, Yuqing Guo, Amir M. Rahmani

**Affiliations:** Department of Computer Science, University of California, Irvine, Irvine, CA, United States; Bouvé College of Health Sciences, Department of Health Sciences, Department of Pharmacy and Health System Sciences, Northeastern University; Sue & Bill Gross School of Nursing, University of California, Irvine, Irvine, CA, United States

**Keywords:** reinforcement learning, physical activity, contextual bandit, recommendation system, intervention, mHealth system, active learning

## Abstract

**Background:** There are indisputable health benefits to physical activity (PA). By collecting and displaying individual exercise behaviors via wearable trackers, the Internet of Things (IoT) and mobile health (mHealth) have made it possible to correlate users’ physiological data and daily activity information with their fitness requirements.

**Objective:** This study aimed to recommend personalized exercise to non-pregnant subjects to increase their physical activity level.

**Methods:** We developed smartphone and smartwatch applications to collect, monitor, and recommend exercises using a contextual multi-arm bandit framework. Twenty female college students were recruited to test this mHealth exercise program.

**Results:** Our findings indicated an increase in daily exercise duration (*P* < .001), with average satisfaction scores for the walking and recommendation system components of 4.31 (0.60) and 3.69 (0.95), respectively, on a scale of 1 to 5. In addition, participants’ confidence in their capacity to complete the suggested walking exercises safely and the study’s ability to satisfy their needs for physical activity both received average scores of over 4.

**Conclusions:** A new era of mHealth systems has been ushered in by developments in the Internet of Things and wearable devices. Personalization of physical activity recommendations using such wearables has the potential to improve user engagement and performance. In this paper, we presented an exercise recommendation system based on reinforcement learning that uses biomarkers and the user’s context to recommend a unique walking exercise that enhances the user’s aerobic capacity.

## Introduction

Physical activity (PA) has undeniable health advantages. The World Health Organization (WHO) and a growing number of national governments worldwide have developed public health-oriented PA recommendations in response to the relevance of physical inactivity as a risk factor for chronic illnesses and early death [24].

The Internet of Things (IoT) and mobile health (mHealth) have made it feasible to link users’ physiological data and daily exercise information with their fitness demands by collecting and visualizing individual exercise activities via wearable trackers [13].

Despite the great potential of mHealth services in free-living conditions, the evaluation of exercise recommendations and optimization is challenging. The majority of these mHealth services suffer from non-personalization and having fixed activity suggestions for all the users, which may result in fixed physical activity [27]. Such an inefficiency might have different underlying causes, as described in an earlier study [13].

Several systematic studies [11, 12, 18] investigated the effectiveness of mHealth in physical fitness and interventions, as well as the need for mHealth technology to promote physical health. Just-in-time adaptive models [13, 14], reinforcement learning-based models [15, 17, 8], neural network-based models [19] are such instances of mHealth designs. Although these studies claim personalization, they are limited in their ability to monitor real-time heart rate as well as the intensity during the exercise. Machine learning models can be practical for extracting the patterns of entire data sets, and they work best when there is enough data; however, in a real-time system at the beginning of a study, the sparsity of the data makes the prediction task challenging. Therefore, Reinforcement Learning (RL) gained enormous attention as they learned the policies by observing the rewards of different user actions. These RL algorithms have been applied in a wide range of healthcare applications [15,16,17].

In this study, we designed the first end-to-end closed-loop physical activity recommendation system in the wild using personalized real-time heart-rate (HR) monitoring with a proof-of-concept study. We designed and implemented a contextual bandit framework as an adaptive (active) learning approach to recommend exercises based on personalized biofeedback, exercise intensity, and overcome the cold start problem at the beginning of the study. Our result showed an increase in daily exercise duration (*P* < .001) and walking and recommendation system components had average satisfaction scores of 4.31 (0.60) and 3.69 (0.95), respectively, on a scale of 1 to 5. In addition, participants’ confidence in their capacity to do the suggested walking exercises safely and the study’s ability to satisfy their needs for physical activity both received average scores of over 4.

To summarize, our main contributions include:

1. We designed and implemented a closed-loop mHealth system with personalized exercise recommendations. The users can interact with different interfaces using smartphone and smartwatch applications.
2. The system leverages the user’s real-time heart rate and exercise intensity as inputs for the system.
3. We conducted a 12-week study with 12 active participants as a proof of concept to demonstrate the functionality and effectiveness of the system.

## Methods

### Recruitment

Study information was sent to undergraduate and graduate students through the School of Nursing email list in November 2021. There were 52 students who showed interest in filling out the screening form. Twenty female participants were recruited for the study. Participants were eligible to enroll in the study based on the following criteria: females 18-40 years old, healthy without any medical conditions preventing them engaging in exercise, spoke English, owned a smartphone (iPhone or Android compatible with the UNITE app), and agreed to wear the smart devices as well as complete the recommended exercise activities and submit surveys related to their experiences. Participants’ characteristics are presented in Table 1.

**Table 1.**
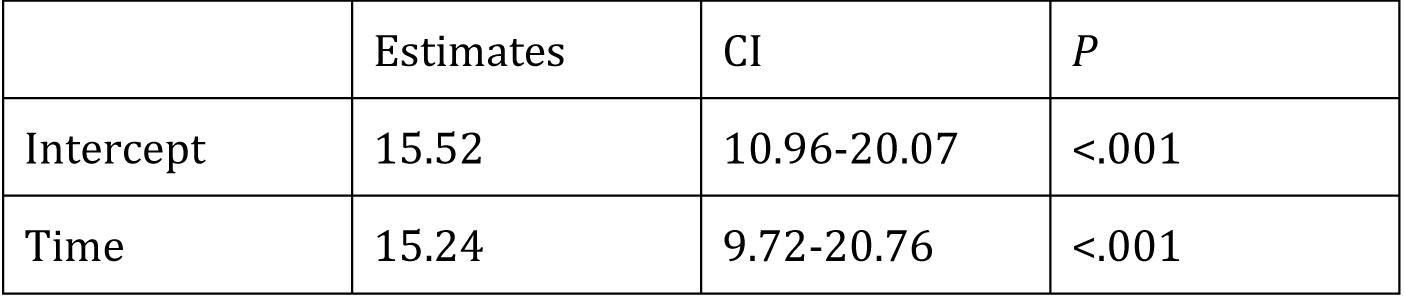
Summary of fixed effects of the HLM fitted to the exercise performed duration.

|  | Estimates | CI | *P* |
| --- | --- | --- | --- |
| Intercept | 15.52 | 10.96-20.07 | <.001 |
| Time | 15.24 | 9.72-20.76 | <.001 |

**Table 2.** Summary of random effects of the HLM fitted to the exercise performed duration.

|  | Standard deviation |
| --- | --- |
| Residual | 7.88 |
| Intercept | 6.98 |
| Time | 6.43 |

### Table 1. Subject Characteristics (n=20)

- ○ Level 1: 15-minute walk three days a week: 9 subjects
- ○ Level 2: 20-minute walk three days a week: 10 subjects

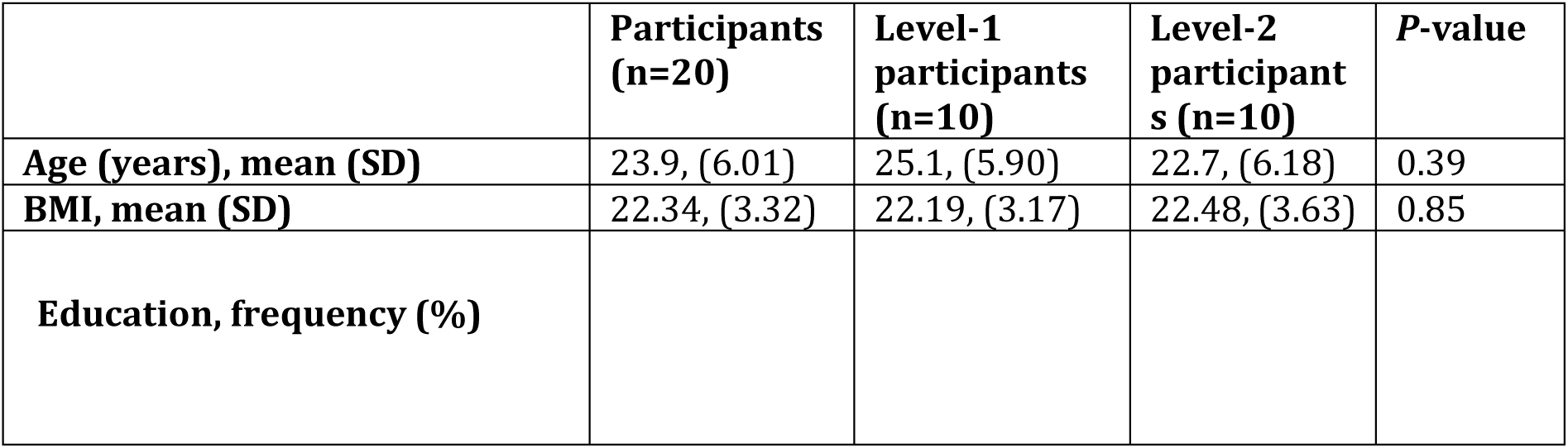
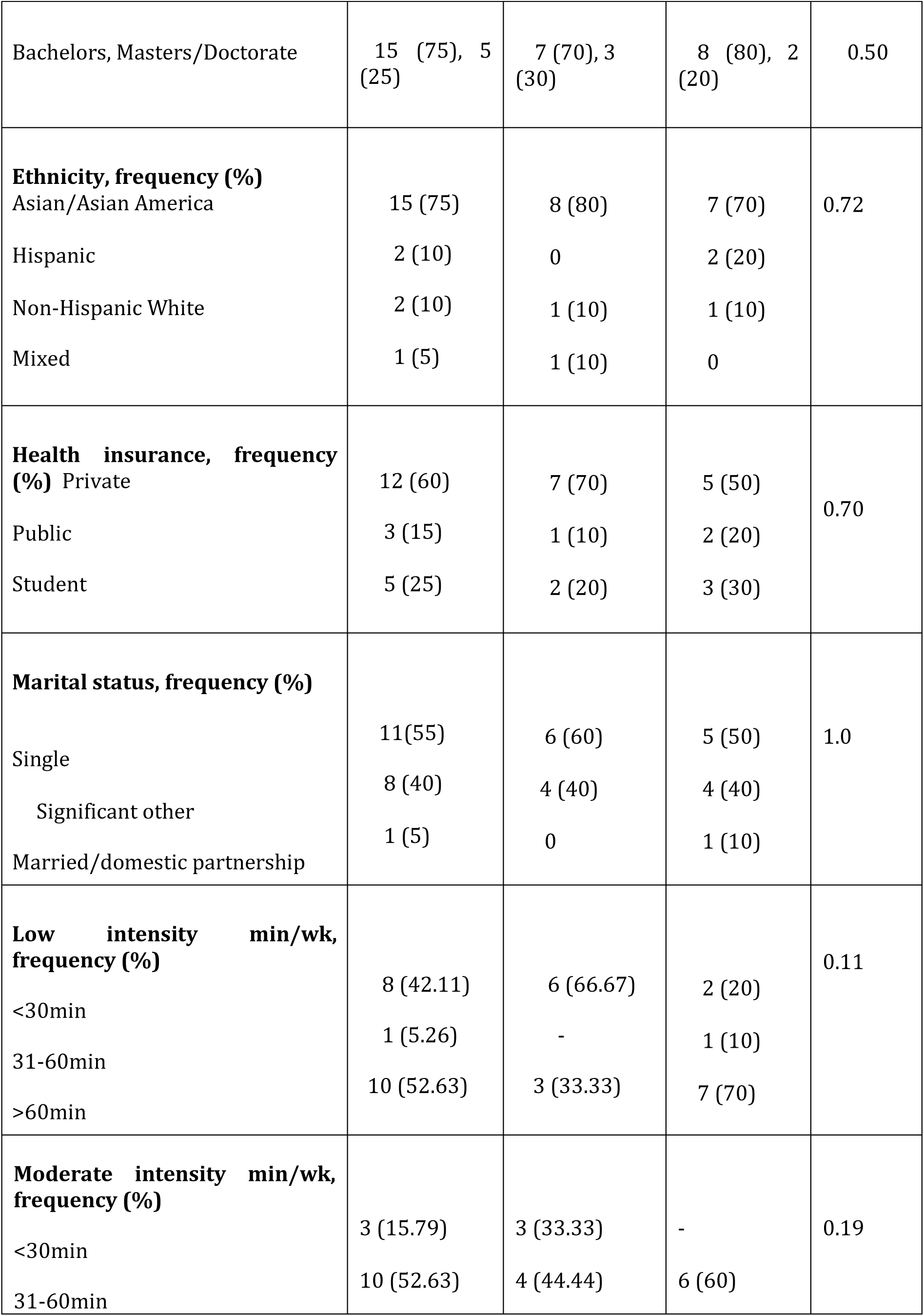
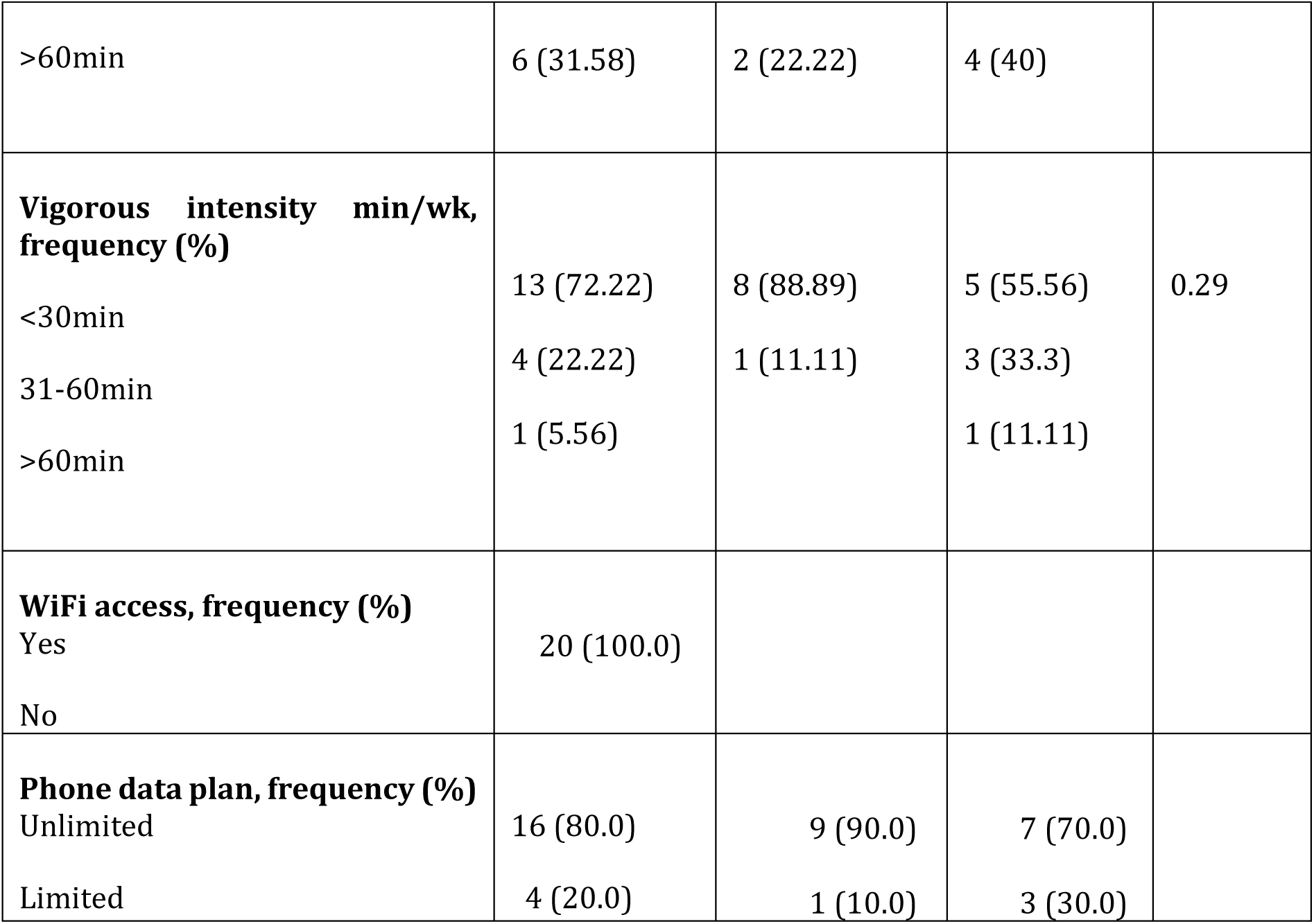

### Study Procedures

The study was approved by the University Institutional Review Board (IRB). The trained research assistant consented twenty participants and provided them a Study Information Sheet with detailed explanation. All consented participants received the REDCap link to complete baseline demographic and exercise history questions. In addition, the research team supported them to properly install and connect the wearable smart devices to the smartphone; a virtual group session was held to provide an overview of the study, explain the recommendation system as well as answer any questions participants had. Participants were first assigned to one of two exercise activity levels depending on their exercise history. Subsequently, participants completed weekly walking activities with the recommended frequency and duration determined by their heart rate and self-reported soreness and discomforts using the UNITE app surveys. Four participants withdrew in the middle of the study due to time constraints. Sixteen completed the 12-week personalized walking program and filled out the exit surveys assessing participants’ user experiences on Likert scale 1 to 5.

### Exercise organization

Ideally, human subjects should get at least 150 minutes of moderate-intensity aerobic activity every week according to WHO recommendations [29, 30] This moderate-intensity activity can be relative to an individual’s fitness level [1]; such movements can raise the heart rate to a certain level. Aerobic activity includes a large portion of muscle movement in a rhythmic way [28].

Borg test, as well as talk test, are two classical methods for exercise intensity detection. The Borg test contains a numbered list of values that a subject reports in response to the intensity level of an exercise. The drawback of such tests could be generalizability. In addition, such tests are subjective and cannot accurately capture an exercise’s intensity. Similarly, as long as an individual can follow up on a conversation while exercising, she is not in the moderate-intensity zone [2].

Another exercise intensity detection method relies on heart rate (HR) values during exercise activity. This method is more objective and robust. Maximum heart rate (HRmax) is commonly used to identify different HR zones. The exact value of HRmax can be identified while performing a VO2max test for individuals; however, this method is not feasible in free-living conditions and daily life [4]. Hence, we used the most common equation for HRmax, which is 220 – age [3]. Using HRmax, different guidelines suggest HR ranges for moderate-intensity activity. We used the American College of Sports Medicine (ACSM) recommendation for this study. According to ACSM, any physical activity that increases the HR to 63% of the HRmax is labeled as light intensity exercise. Activities ranging from 64% to 76% can be classified as moderate-intensity exercises, and anything above 77% would be considered vigorous exercise [3].

Literature shows that the lack of time is the main reason for not performing physical activities [24]. Studies indicate that even 30 minutes of movement per week can improve health [24]. Hence, since we had a healthy younger population cohort, we chose 15 and 20 minutes as the baseline for level 1 and 2 groups, respectively. In addition, for inactive people, rather than meeting the physical activity guidelines (150 minutes of moderate-intensity), the global action plan is to start with a baseline and perform small incremental increases in physical activity [25, 26]. Hence, we designed an incremental plan for our exercise recommendation. Figure 1 illustrates such a plan.

**Figure 1.**
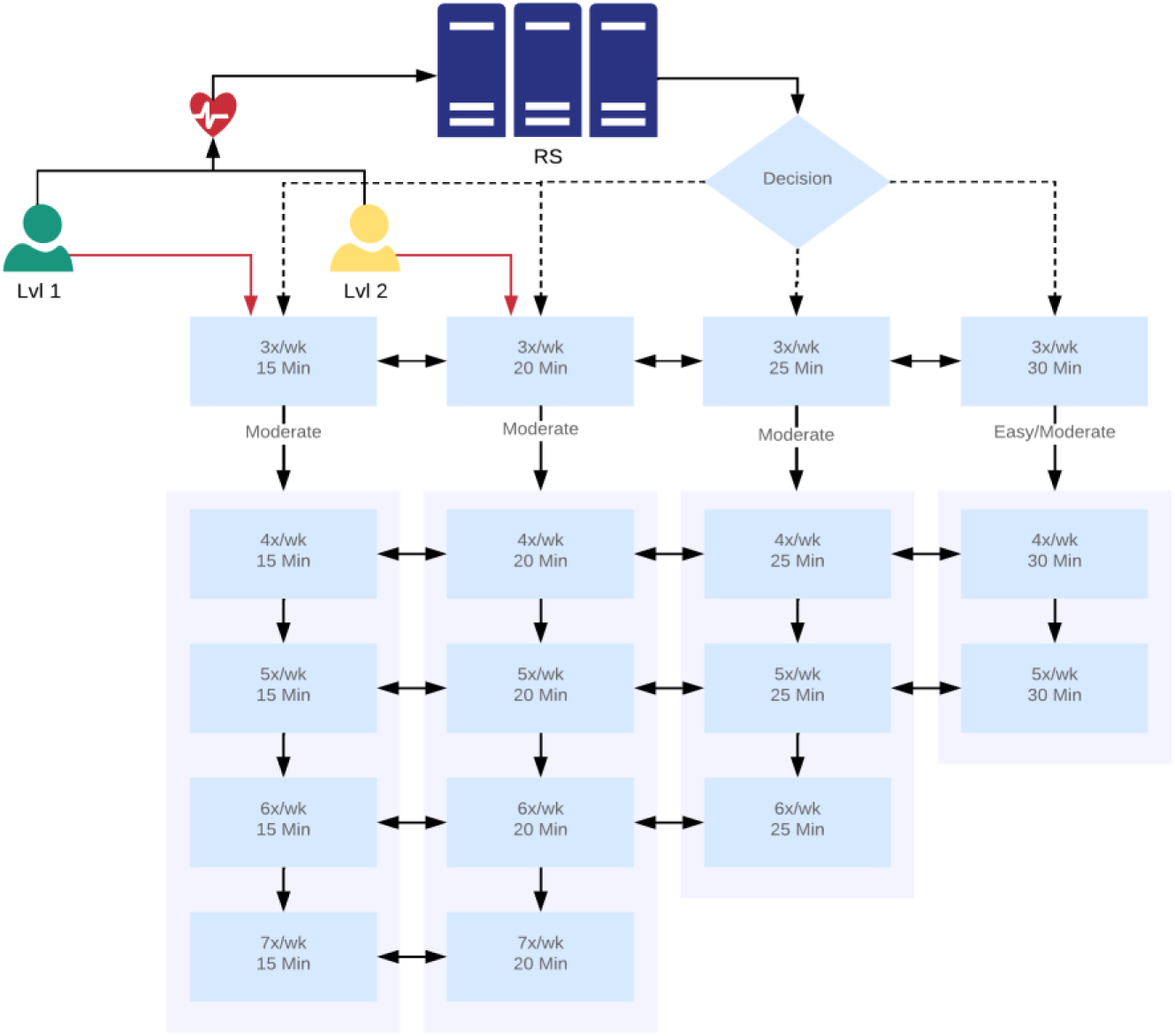
Exercise durations setup.

### Recommendation System Design

#### UNITE mobile smart phone and Samsung watch applications

For this study, we developed two sets of applications and services. The first group contained the applications for users’ mobile devices (i.e., iPhone and Android platforms), which were used to record questionnaires and show the recommendations. The second group consisted of applications developed for the user’s smartwatch (i.e., Samsung active 2) to record the user’s biomarkers and real-time activities. Figure 2 shows an overview of the user interface of these applications.

**Figure 2.**
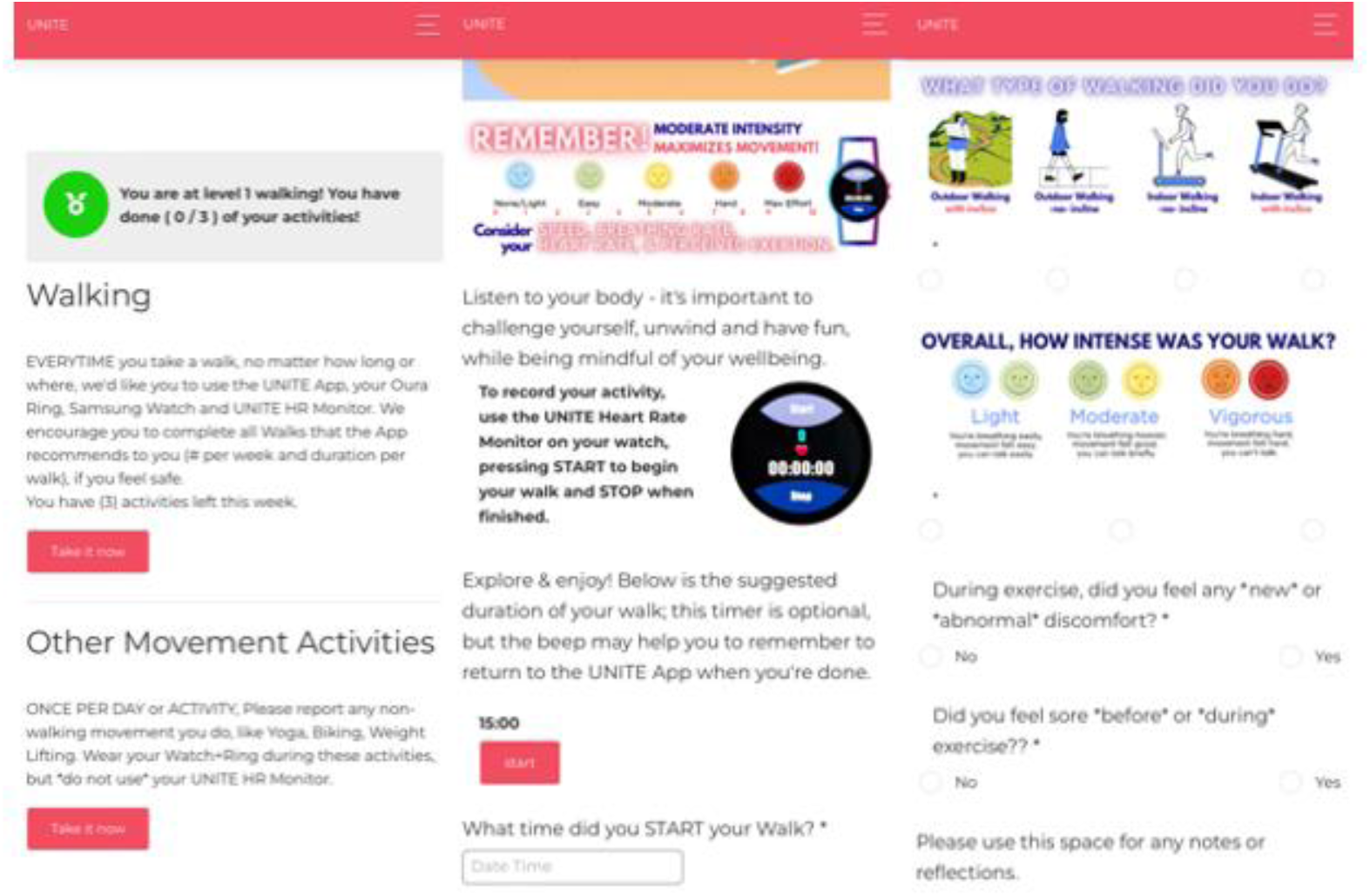
An overview of the developed applications for smartphones and Samsung smartwatch.

#### ZotCare Dashboard

The ZotCare platform [31] has been developed by our group at the University of California Irvine and can provide services for real-time data collection. This platform’s dashboard is designed so experts without programming knowledge can create and manage study parameters, including questionnaires, mobile notifications, user sensor data, and user profiles, in a privacy-preserved form. Figure 3 demonstrates a sample page of such a dashboard for our study.

**Figure 3.**
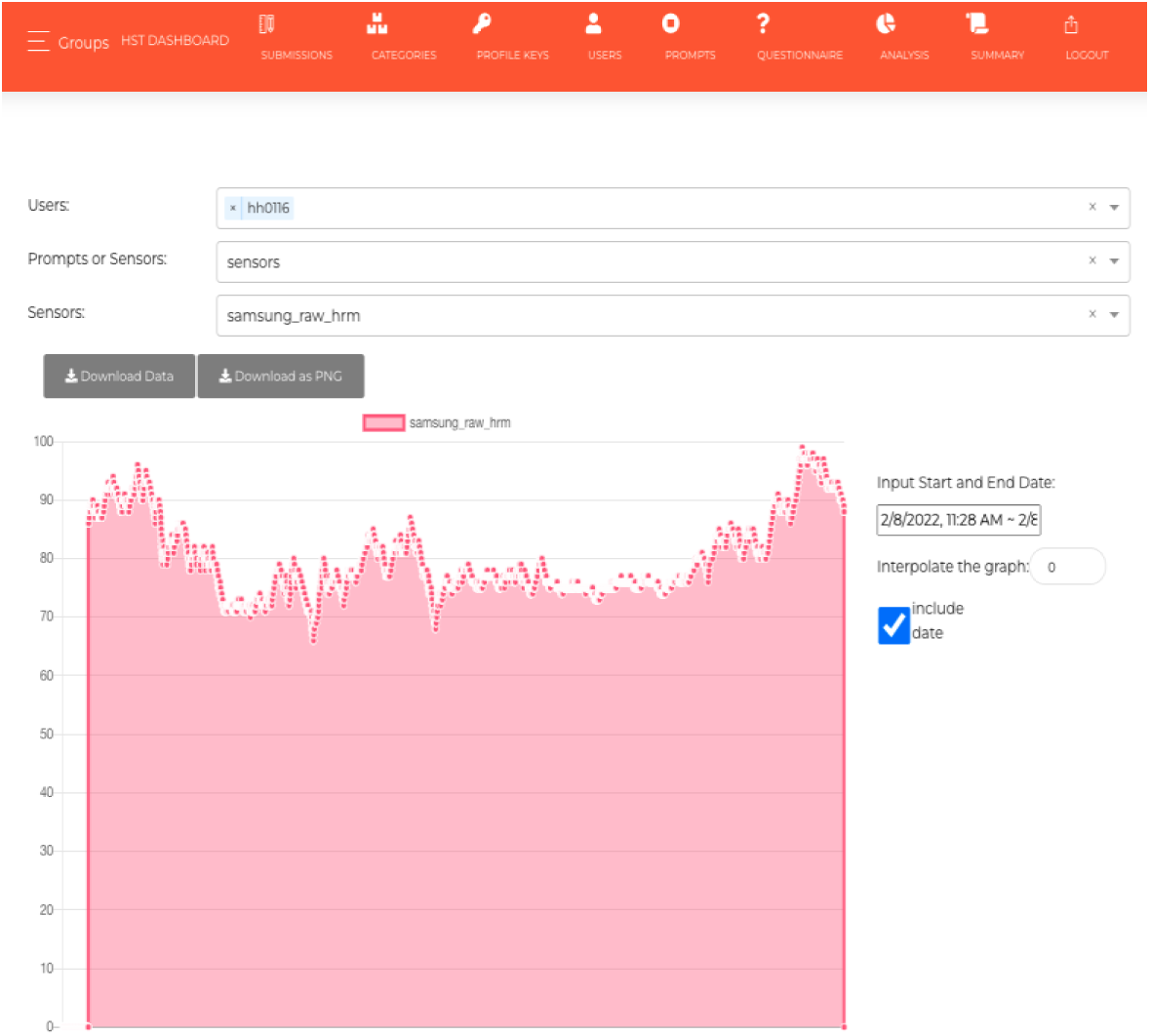
An example page of the ZotCare dashboard.

#### Contextual-bandit Problem

The multi-armed bandit (MAB) algorithms have recently become popular in different domains, including healthcare and recommendation systems. These models are used to maximize the total payout of an agent who is interacting with an environment. At each timestamp *t*, the model selects an action based on a policy, delivers that to the patient and monitors the reward regardless of the user’s context [23].

In exercise recommendations, it is possible to provide a list of suggestions according to their intensity and caloric benefit. Currently, these personalizations are provided only by human health coaches. However, such a method does not capture each individual’s dynamic characteristics, which can change person-to-person or/and over time. Furthermore, the limitation of the data makes it difficult to have perfect predictions, and the cold-start problem is a challenge for such a task. In many applications, there is rich information regarding each action, so having features for every (context, action) pair rather than features associated only with context and shared across all actions would be the more profitable approach.

The Contextual multi-armed bandit can address the challenges mentioned earlier. In the contextual multi-armed bandit (CMAB) problem, a learning model monitors the user’s current context, recommends an action (i.e., exercise in our context), and observes the reward of the recommended action. The objective of such a model is to minimize the cumulative sum of losses with respect to the context. The benefit of the contextual bandit learner compared to the regular multi-armed bandit is the context which makes CMAB personalized and an ideal alternative for dynamic environments. In healthcare, CMAB enables precision medicine to leverage suitable actions for individuals instead of similar actions for multiple patients [4].

There are different algorithms to explore the finite set of actions, and for this study, we used the Epsilon-Greedy approach. In this approach, the model explores the arms (exercises) with a probability of epsilon (0.05 in our case) and with a probability of 1-epsilon exploits the known actions. There are two main reasons for the exploration trade-off. First, to examine new actions, and second, in mobile health setups, although interventions might tend to have positive feedback on the selected action, they can have a negative impact on future rewards because of the user’s habituation [8].

#### Notations and cost functions

In the contextual bandit problem, the environment generates a pair (*x*_*t*_, *l*_*t*_) at time t such that *x*_*t*_ is a context vector and *l*_*t*_ is the loss vector. The learner chooses an action *a*_*t*_and observes the loss of the corresponding action. The objective is to sustain a small cumulative regret

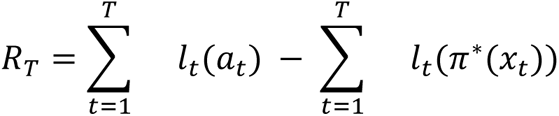

over time [6]. In this equation, *π*^∗^ ∈ *argmin*_*π*∈*ϕ*_*E*_(*x*,*l*)_[*l*(*π*(*x*))] is the optimal policy with *ϕ* being the set of policies.

#### Optimization

There are different methods to solve the optimization problem. We used the regression with importance weights *w*_*t*_ > 0 introduced by [6].

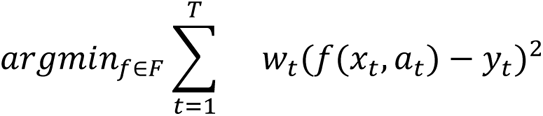

The goal is to find a *f* from a class of regressor functions *F* to predict a cost *y*_*t*_ having the context *x*_*t*_ and action *a*_*t*_.

There are different policy evaluation approaches. The direct method is the most straightforward approach, which maps context and actions to the rewards using a regression model [5]. Since most of the directed methods suffer from high bias, in this study, we used Importance Weighted Regression to reduce the variance and bias of the estimator [6]. This method optimizes a regressor *f̂* to find the optimal policy *π̂* (*π̂* = *argmin*_*a*_ *f̂*(*x*, *a*)) which has been used in off-policy learning recommendation scenarios [7].

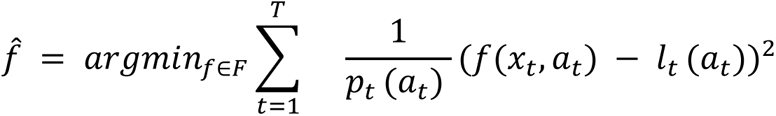

We considered user identifiers, weight, and the number of days from the beginning of the study as the input features (i.e., context). We had access to the sleep features (e.g., total sleep time, resting hr, sleep quality) using the Oura ring; however, due to the manual sync issue, we did not have access to the real-time features. Hence, for this study, we discarded those features for real-time analysis and only performed offline analysis.

The duration of the exercises was considered as the actions for this study. In the case of moderate-intensity duration, we increased the exercise frequency. We trained two different models: one for level 1 subjects and another for the level 2 subjects.

#### Feedback Policy (exercise intensity)

To model the intensity feedback of the exercise, we used the subject’s HR during each exercise. With the help of the ZotCare system [31], we were able to capture semi-real-time data recorded by the Samsung Active 2 smartwatch. Since the smartwatch is prone to noises during recording and data collection, we removed invalid values and used a 10-second moving average filter over recorded non-zero HR data. Later, the exercise reward for the subject *s* was defined as below:

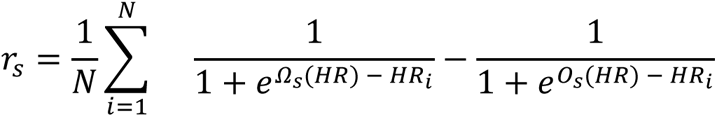

The *Ω* and *O* in this equation represent 64% to 76% of HRmax (i.e., moderate-intensity range) for each subject *s*. *HR*_*i*_reflects the *i*th HR during the exercise. This reward function is convex which meets the requirement for the convexity of the cost function.

The intuition behind this reward function is that for HR values in the moderate-intensity range, the inner part of the summation tends to be close to 1.0. For values outside this range, this term will be close to 0. The average makes this reward function smoother as well as by increasing the number of HR points in the moderate-intensity range, the reward function increases, consequently. Figure 4 depicts the reward function and the corresponding moderate-intensity range for a sample user with *Ω* = 131.3 *bpm* and *O* = 151.5 *bpm*.

**Figure 4.**
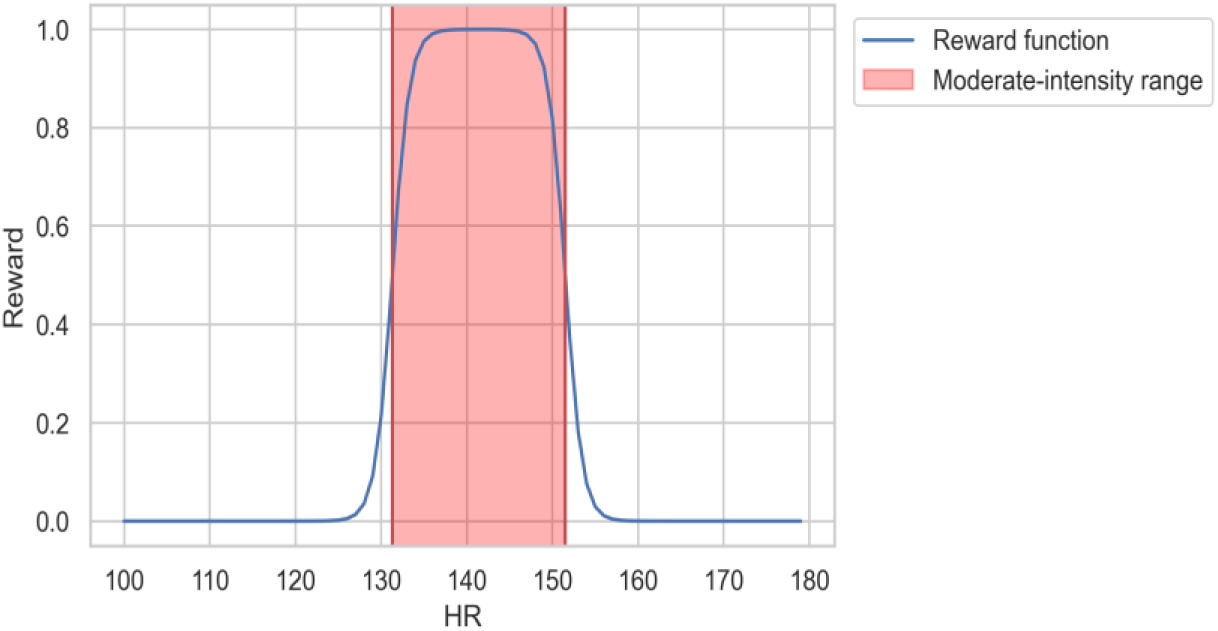
Reward function with respect to the HRs during the exercise.

Figure 5 outlines the entire recommendation system design. Different data sources record the subject’s data and continually send it to the servers using Wifi/Bluetooth/Mobile network connection. The ZotCare system manages the different modalities of data, and the recommendation system pulls different data using its cloud interface. This system makes decisions regarding different alerts and/or updates on exercises.

**Figure 5.**
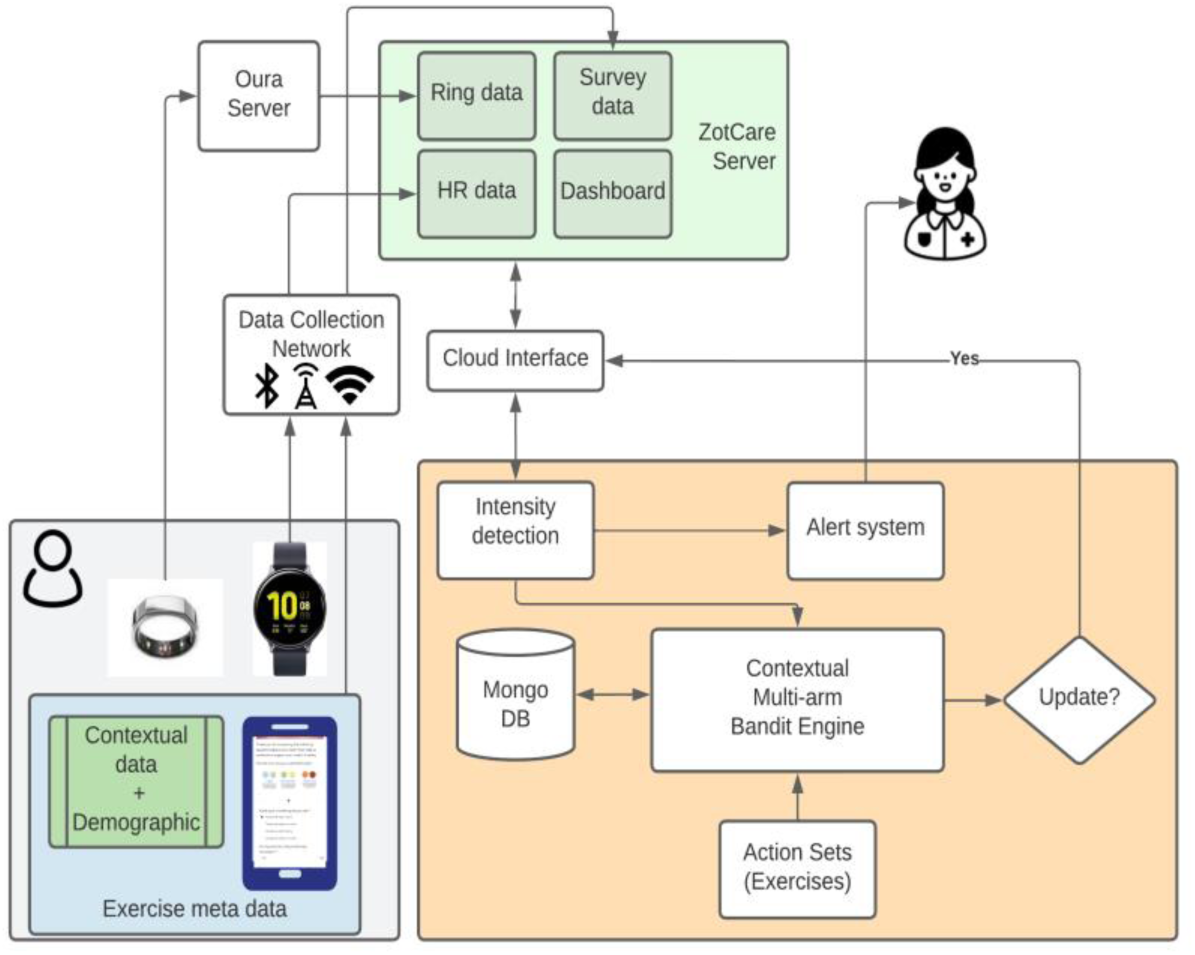
Overview of the system.

#### Safety module

To ensure that the exercises are safe, we designed an alert system notifying the nurse about the outcome of the exercise. The alerts are generated in case the subject’s HR exceeds 90% of the HRmax, she feels new discomfort during the exercise, and/or has soreness due to the exercise. In case of any of the outcomes above, the recommendation system maintained the same workout for one more session.

### Statistical Analysis

#### Data Exclusion

During our study, one subject did not participate in any of the exercises, and two subjects dropped out due to personal reasons. Out of the rest, we had four subjects who were experiencing technical difficulties and/or restarted their watch by mistake. Hence, we lost a large portion of their data. Since we were programming the watches manually, the turnaround time was causing us to lose data. In addition, a couple of these subjects submitted the reports incorrectly, making it challenging to track their physical activities. Although these subjects contributed to the reinforcement learning framework, we excluded them from the offline analyses.

#### Mixed-Effect Analysis

Hierarchical linear mixed (HLM) models (also known as mixed effect models) were exploited to analyze the trends in between- and within-subject variances. We did this analysis using the notation defined and recommended by Raudenbush-Bryk and Bolger-Laurenceau [9, 10]. The dependent variables of interest include duration in light/moderate intensity over time, and exercise duration performed by the subjects.

## Results

### User Statistics

This section explains the trend of both performed and recommended physical exercise for each participant and their submission timelines.

### Improvement in performed exercise duration trends

During 12 weeks of this study, we monitored subjects’ exercise behavior recorded by our application. We only considered those exercises that the subject reported and had HR data available.

Figure 6 shows the average exercise duration with standard error bars for each group (i.e., level 1 and level 2) over the study period. In general, it shows an increasing trend throughout the study. In addition, we utilized mixed effect models to illustrate the within- and between-subject trends over time.

**Figure 6.**
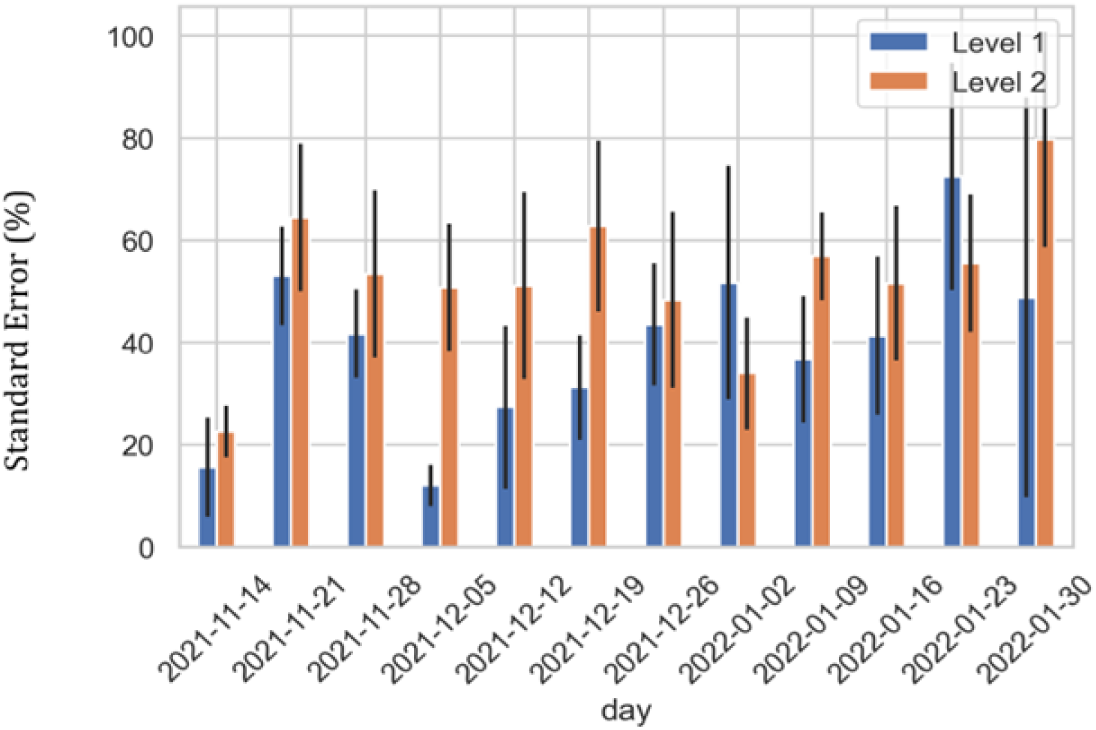
Average weekly duration of walking for each group. The bars represent standard error percentage.

Figure 7 illustrates the trends for each subject. The blue dots show the actual duration that the subject did, and the red ones demonstrate the recommended duration.

**Figure 7.**
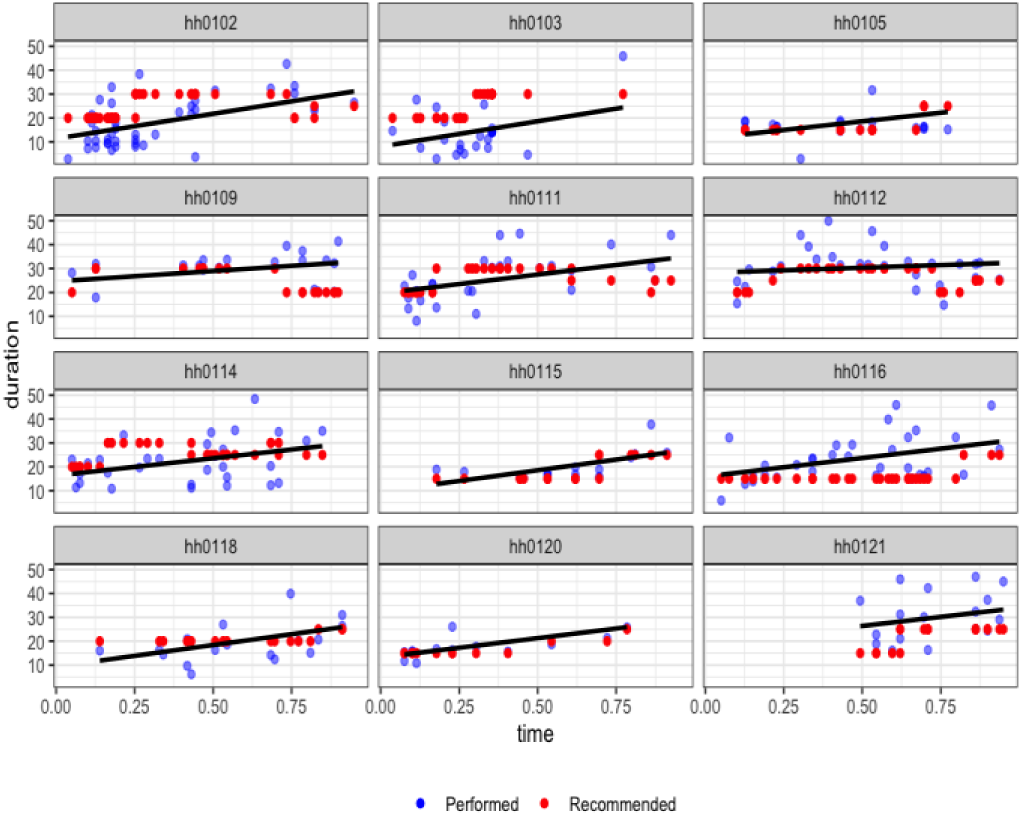
Performed vs. recommended duration of walking per each subject. The line represents the fitted HLM model to each subject.

Tables 1 and 2 summarize the fixed and random effects of such a model. To overcome the exploding gradients effect, we normalized the x-axis (time) before applying the mixed effect model. The general trend for these subjects is significantly increasing (*P* < .001), with an initial intercept of 15.52 minutes of exercise on average (*P* < .001). The correlation of fixed effects is *r*_*f*_ = −0.81, and the correlation of random terms is *r*_*r*_ = −0.84. This is not surprising since subjects starting with a lower initial level tend to reach the goal faster than other groups (i.e., level 2 subjects). Figure 8 illustrates the quantile-quantile plot (Q-Qplot) and normality of the residuals.

**Figure 8.**
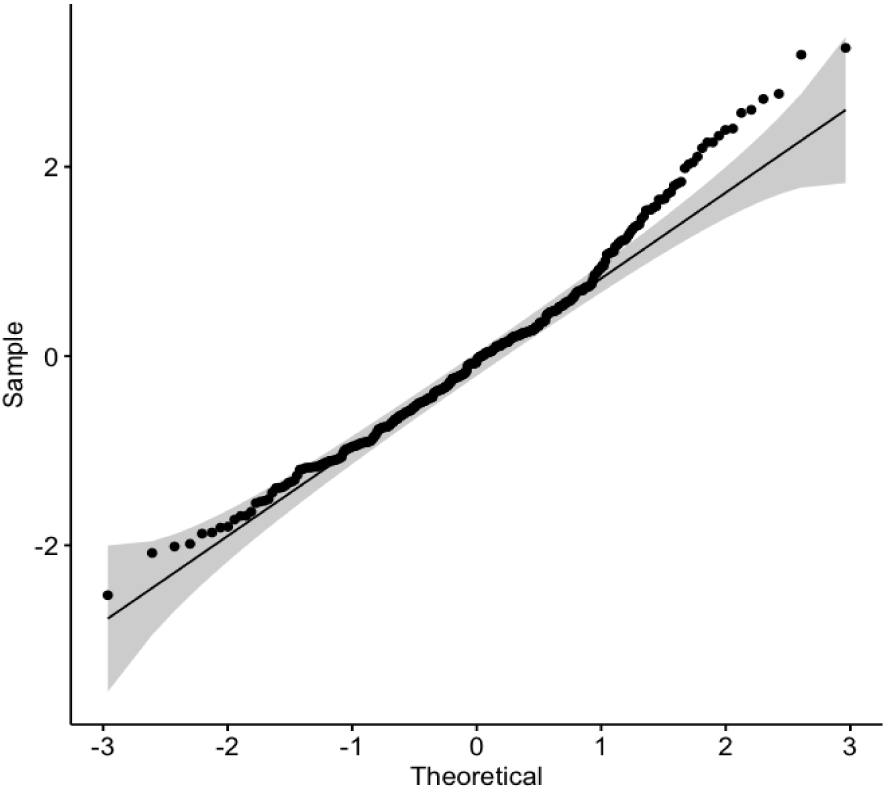
Normal Q-Q plot for normality of the residuals for exercise performed duration.

### Minutes in light- and moderate-intensity

In addition to the exercise total duration, the HLM model was utilized to analyze the trend of non-vigorous activity duration (duration in which the HR is below 77% of the HRmax). Figures 9 and 10 illustrate per subject trend estimation and dot plot, respectively.

**Figure 9.**
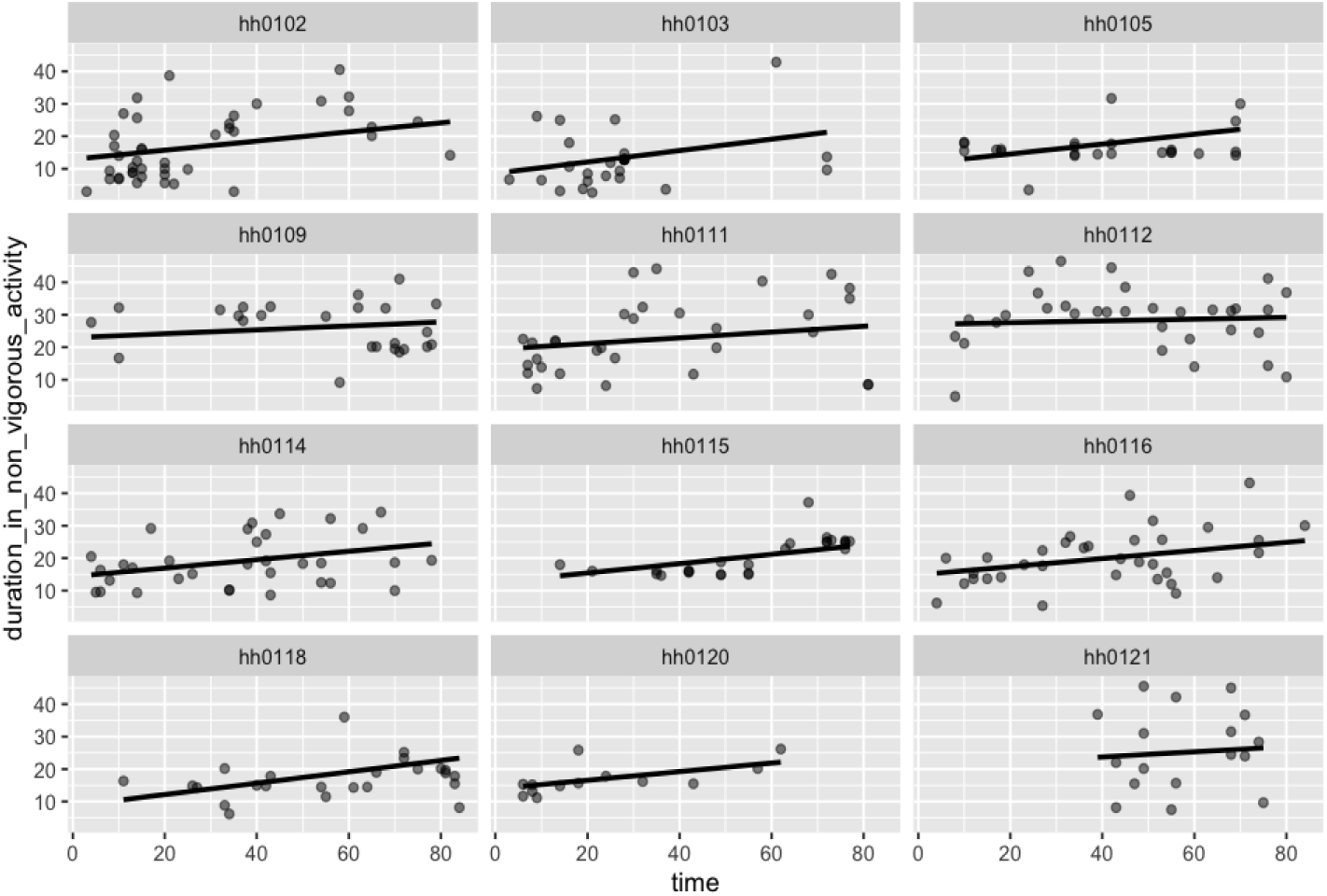
Trends in minutes in light and moderate-intensity exercise over time.

**Figure 10.**
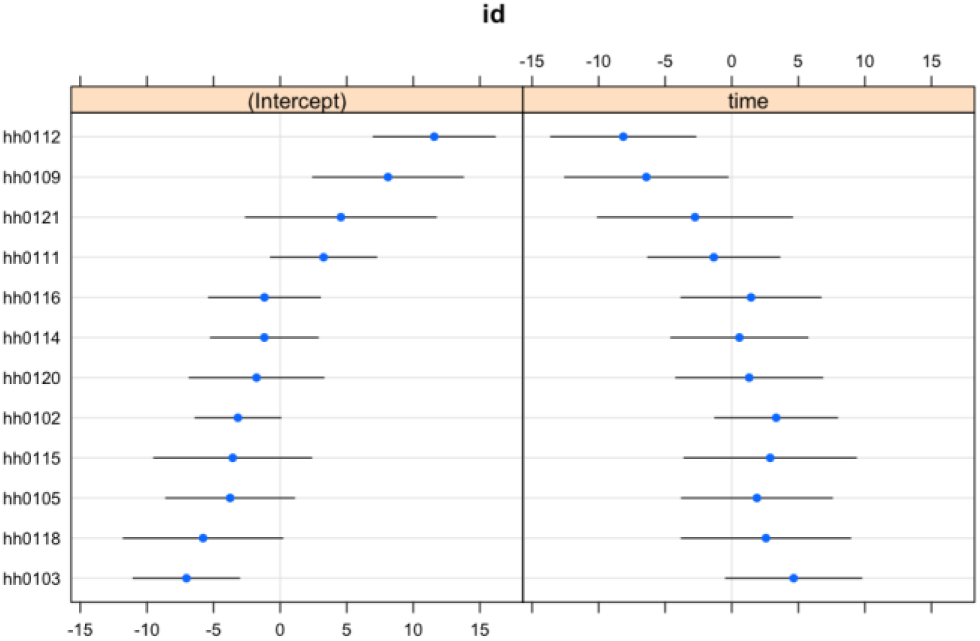
Dot plot visualization of between-subject variations compared to the mean trend.

Tables 3 and 4 show the fixed and random effect for light- and moderate-intensity exercise duration. The general trend significantly increases the corresponding duration (*P* < .001). The effect correlations are *r*_*f*_ = −0.84, and *r*_*r*_ = −0.93. Figure 11 shows the Q-Q plot for the residuals. Subjects hh103 and hh112 have the highest and lowest increase, respectively.

**Figure 11.**
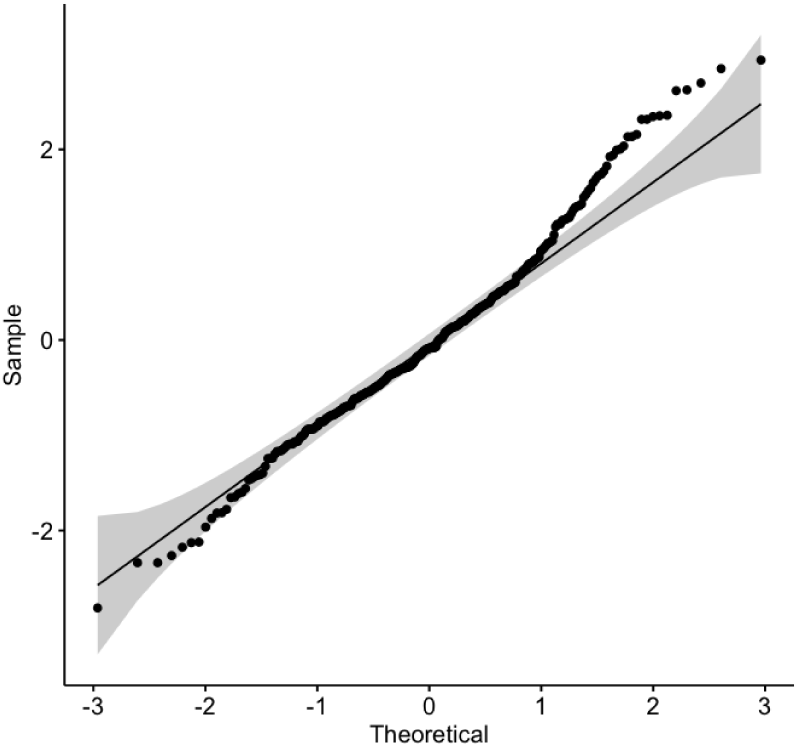
Normal Q-Q plot for normality of the residuals for minutes in light/moderate-intensity exercise.

**Table 3.**
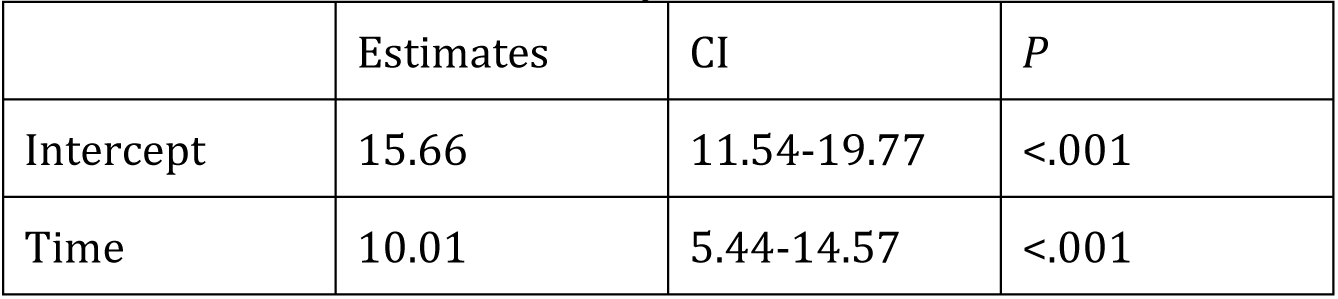
Summary of fixed effects of the HLM fitted to the minutes in light/moderate-intensity exercise.

**Table 4.** Summary of random effects of the HLM fitted to the minutes in light/moderate-intensity exercise.

|  | Standard deviation |
| --- | --- |
| Residual | 8.17 |
| Intercept | 6.25 |
| Time | 4.92 |

### Aggregated weekly performance

To investigate the weekly performance of the users and compare it with the standards and guidelines, we aggregated the exercise execution of the subjects over each week of the study. Figure 12 shows the bar plot of weekly exercise duration for each user regardless of the longitudinal notion of time. Each color represents the exercise minute ranges. Subjects hh116 and hh102 were able to perform more than 150 minutes of weekly exercise.

**Figure 12.**
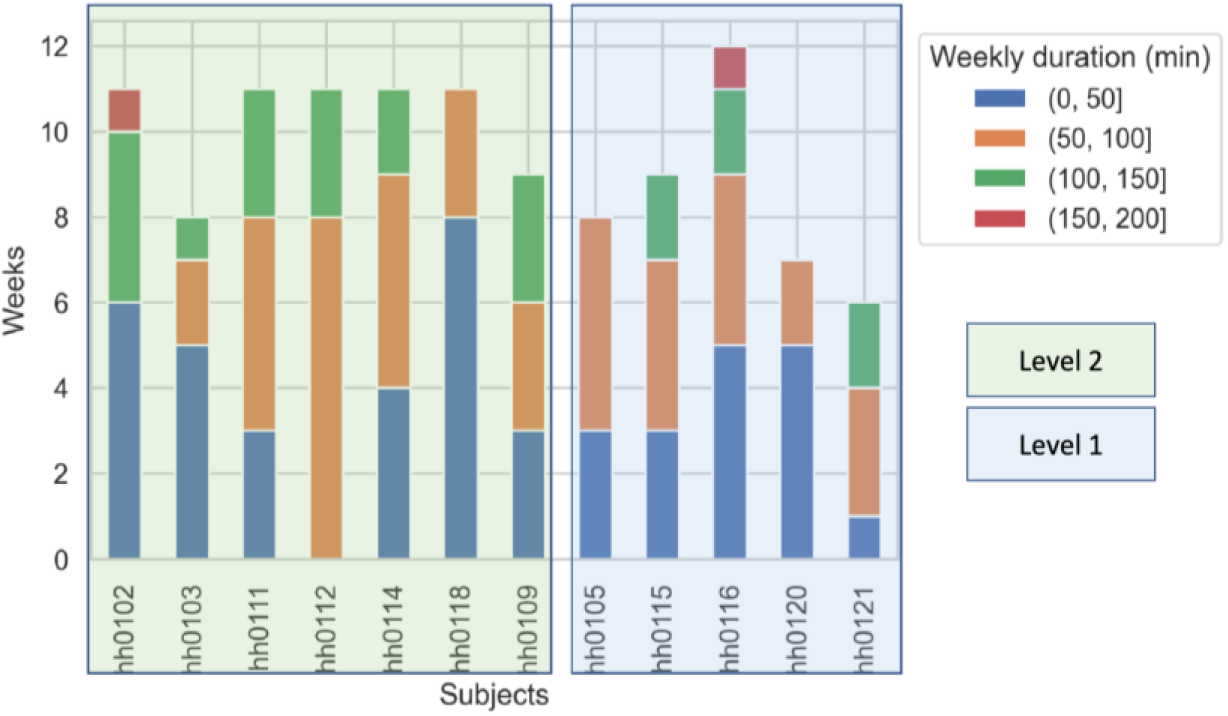
Aggregated number of weekly exercise duration performed by the subjects.

Furthermore, we investigated the weekly exercise trends of the subjects by applying HLM models to the weekly durations over time. Figure 13 indicates such trends for each user, and Tables 5 and 6 summarize the fixed and random effects. The effect correlations are *r*_*f*_ = −0.86, and *r*_*r*_ = −0.85. According to Figures 12 and 13, the results show an increasing trend for subjects; however, for some of the weekly results, the trend is decreasing (e.g., subject hh102). The investigation showed that these subjects tended to submit multiple walking exercises in one day towards the end of the study. There may be several explanations for this behavior, including the decline in user engagement throughout the study discussed by [22]. Since our recommendation system runs daily, only the last exercise performed that day will be considered, which causes the aforementioned issue in the exercise monitoring system.

**Figure 13.**
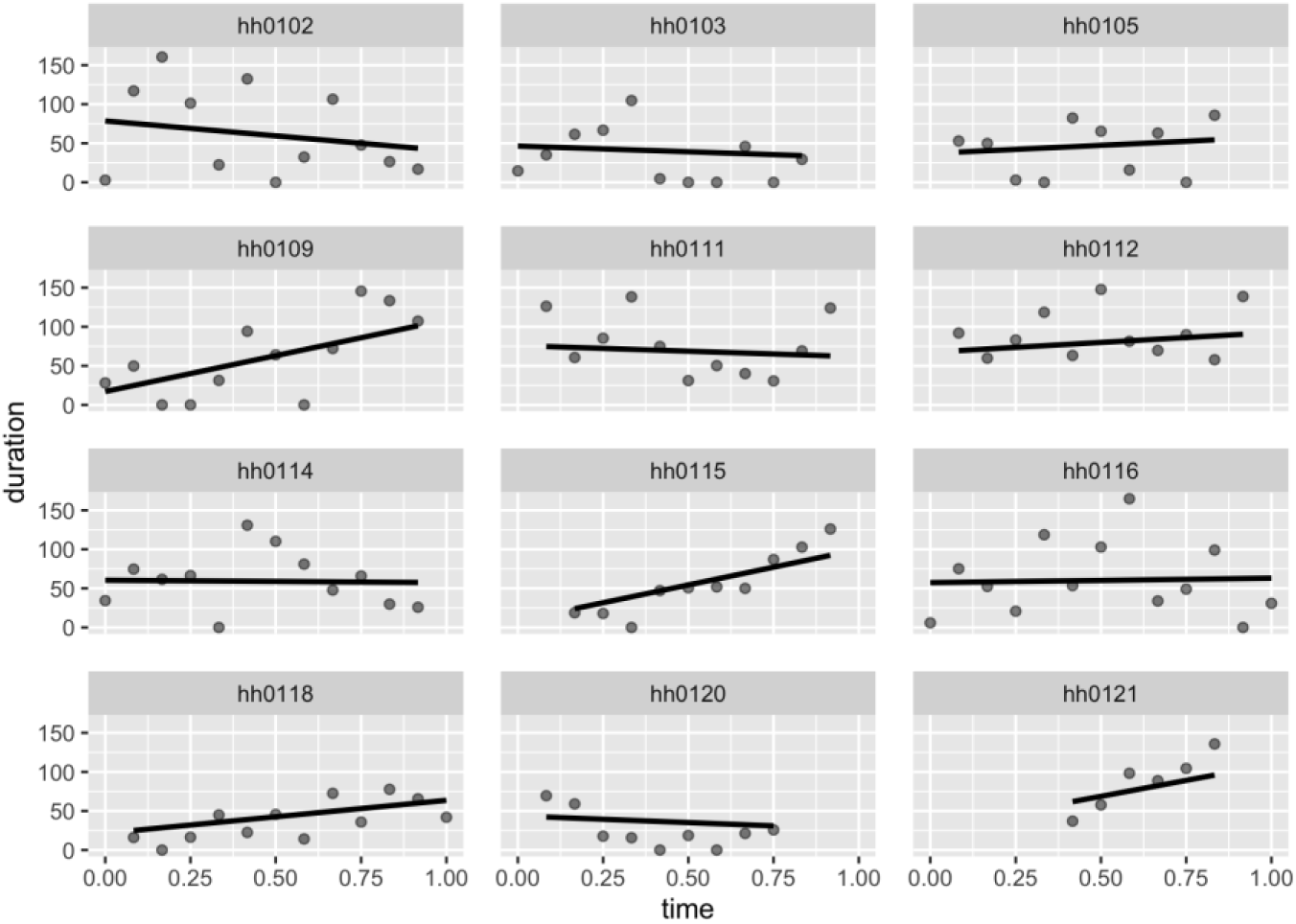
Trends in weekly exercise duration.

Tables 5 and 6 show the fixed and random effect for light- and moderate-intensity exercise duration. Figure 14 shows the Q-Q plot for the residuals.

**Table 5.** Summary of fixed effects of the HLM fitted to the weekly exercise duration.

|  | Estimates | CI | *P* |
| --- | --- | --- | --- |
| Intercept | 45.19 | 23.16-67.21 | <.001 |
| Time | 22.57 | -18.19-63.34 | .27 |

**Table 6.** Summary of random effects of the HLM fitted to the weekly exercise duration.

|  | Standard deviation |
| --- | --- |
| Residual | 36.81 |
| Intercept | 30.15 |
| Time | 56.95 |

**Figure 14.**
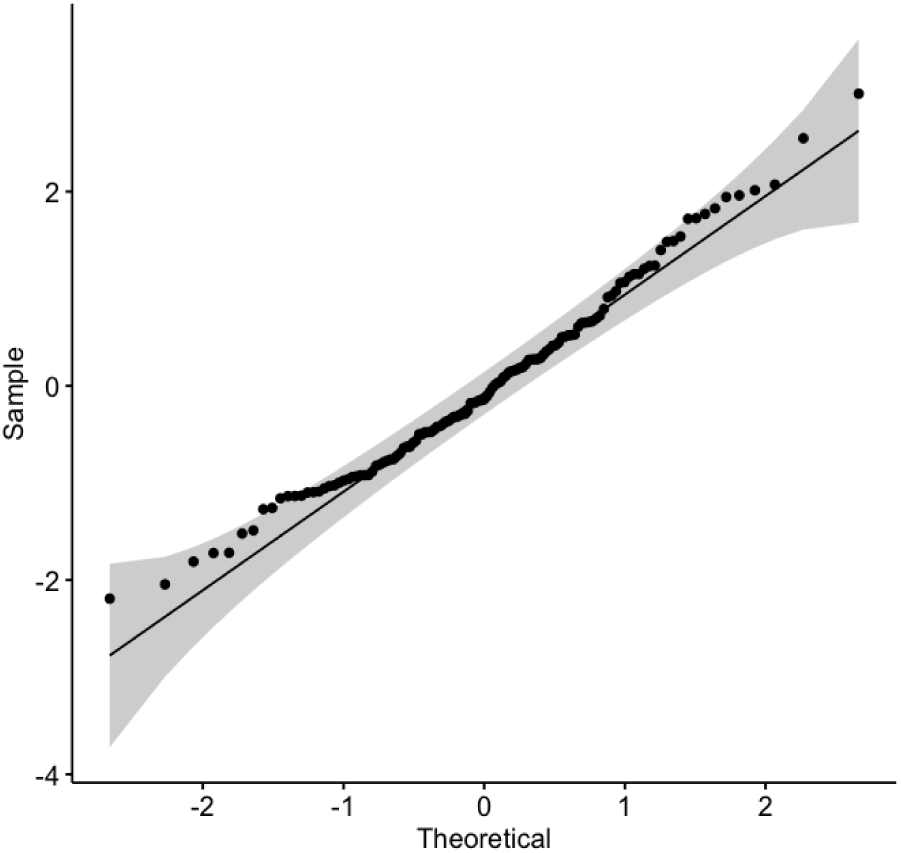
Normal Q-Q plot for normality of the residuals for the weekly exercise duration.

### Recommended exercises

#### Reward analysis

The average cumulative reward has been monitored to evaluate the recommendation system’s performance. In the beginning, since the learner does not know about the subjects, the reward is zero. However, the performance improves as the model explores/exploits different actions given the current context. Figures 15 and 16 show the average reward throughout the exercises being done by each group (i.e., levels 1 and 2). Since two different models were used for the level 1 group and the level 2 group, we evaluated them separately. Although the models differed, the overall cumulative average reward follows the same pattern for both groups. Even though the patterns are the same and increasing, the model for level 2 subjects performed better due to access to a more significant portion of data (about twice as level 1 exercise data). Since the nature of our design is similar to active learning techniques, providing more data increases the system’s performance.

**Figure 15.**
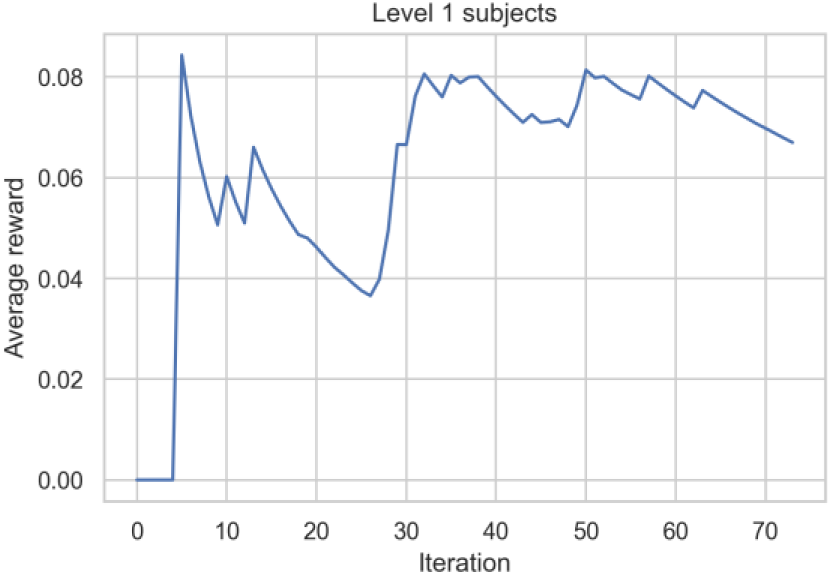
average reward throughout the exercises for group level 1.

**Figure 16.**
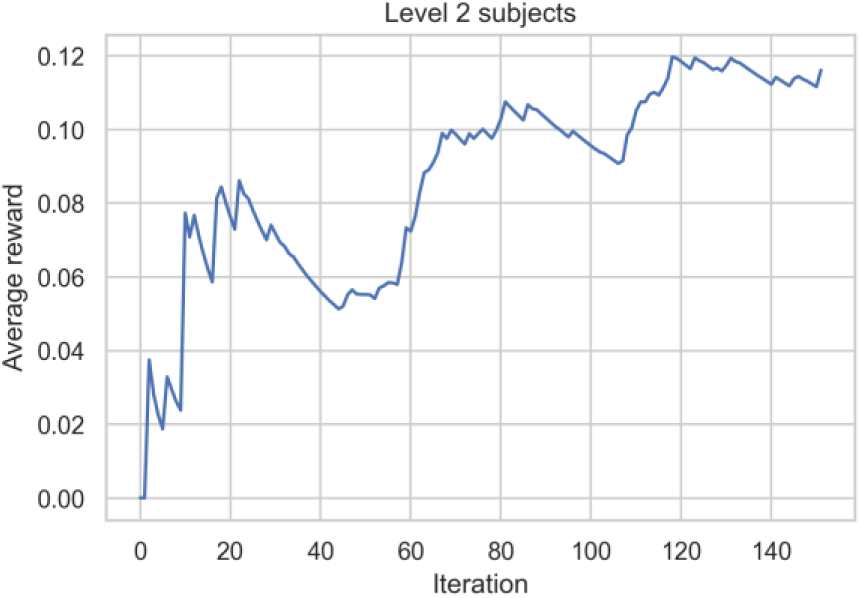
average reward throughout the exercises for group level 2.

### Participants feedbacks (exit survey)

**Exit survey (n=16)**

At the end of the study, exit surveys were administered to participants to understand their user experiences in order to improve the design and implementation of future studies. Overall, 87.5% of participants reported that the exercise recommendation system met their physical activity needs; 75% of participants rated their enjoyment using the smart watch and 100% enjoyed personalized HH walking recommendations).

## Discussion

### Principal Results

To the best of our knowledge, this study is the first end-to-end closed-loop physical activity recommendation system in the wild using personalized real-time HR monitoring with a proof-of-concept. Our proposed model showed an increase in the daily exercise duration of the users which can be useful in getting inactive individuals to perform some physical activity with a comfortable intensity level.

### Limitations

In this study, we had multiple apps designed to capture the data. Users had to manually start the HR monitoring app on their smartwatch, which decreased the convenience of our design. Due to this fact, some users forgot to record their physical activity; hence, retrieving such exercise data was challenging. In addition, we implemented both the Bluetooth and WiFi connection methods in our design. Due to the lower bandwidth of the Bluetooth connection, we experienced some delays in receiving the exercise data.

Moreover, our study had a cold-start problem which caused some initial uncertainty. A pre-trained model over a baseline period could help make more satisfactory predictions. Similarly, the prediction could drastically improve by increasing the duration of the study; however, studies showed that the users’ engagement with the system decreases over time [22].

### Comparison with Prior Work

Different systematic reviews studied the effectiveness of mHealth in physical fitness and interventions and the need for mHealth technologies to improve physical health [11, 12, 18]. Through mHealth technologies, it is now possible to implement just-in-time adaptive interventions [13, 14]. Yom-Tov et al. [15] used a reinforcement learning approach on patients with diabetes to motivate them to increase their physical activity using encouraging messages. The results showed an improvement in the number of successful intervention messages. Saponaro et al. [17] performed a similar study with healthy subjects, capturing different contextual features to evaluate the effectiveness of nudges in free-living conditions. Rabbi et al. [16] designed a recommendation system to generate health feedback based on physical activity and the log of foods. Author in [8] designed a reinforcement learning algorithm to recommend a treatment policy (activity) to increase the step counts. Although they evaluated their algorithm’s performance using different simulations, the algorithm was not tested in the wild. Mahyari et al. [19] designed a model consisting of two inter-connected recurrent neural networks (RNNs) to suggest a new exercise based on the historical sequence of performed exercises. They extracted features from the name of exercises using word2vec and natural language processing techniques. Furthermore, other studies used a contextual bandits recommender algorithm to improve emotion regulation, demonstrating that context is essential for effective emotion regulation [20, 21]. Despite the fact that these studies claim personalization, they suffer from the inability to monitor real-time heart rate during physical activity, as well as the intensity of the physical activity.

## Conclusions

Advances in mHealth and Internet-of-Thing have created a new health monitoring and management system era. Nowadays, a general model cannot be used for interventions with different populations, and there is a need for personalized policies and model buildings. Physical activity needs attention since the human body functionally changes person-to-person and over time. Personalization in physical activity recommendations could potentially improve the user’s performance as well as his/her engagement. In this study, we proposed a reinforcement learning-based exercise recommendation system that utilizes a person’s biomarkers and a context to suggest a new walking exercise that maximizes the user’s aerobic capacity. We showed that this system works in an active learning environment. As a future direction, we are designing such a system for pregnant women, and the reward module works with heart rate reserved since pregnant women’s HR norms change during pregnancy. In addition to the features mentioned in this study, we are utilizing other context features such as sleep quality for the prediction task.

## Data Availability

All data produced in the present study are available upon reasonable request to the authors

## Acknowledgments

This research was funded in part by the Academy of Finland through the SLIM Project under grant numbers 316810 and 316811 and in part by the U.S. National Science Foundation (NSF) through the UNITE Project under grant number SCC CNS-1831918.

## Conflicts of Interest

None declared.

## Abbreviations

PA: Physical activity
HR: Heart rate
ACSM: American college of sports medicine
MAB: Multi-armed bandit
CMAB: Contextual multi-armed bandit
HLM: Hierarchical linear mixed

## Appendix 1

PA intensity: low 1 (<=30min), moderate 2 (31-60min), vigorous 3 (>60min) Low PA intensity for 20 subjects

| low_in | Freq. | Percent | Cum. |
| --- | --- | --- | --- |
| 1 | 8 | 42.11 | 42.11 |
| 2 | 1 | 5.26 | 47.37 |
| 3 | 10 | 52.63 | 100.00 |
| Total | 19 | 100.00 |  |

Moderate PA intensity for 20 subjects

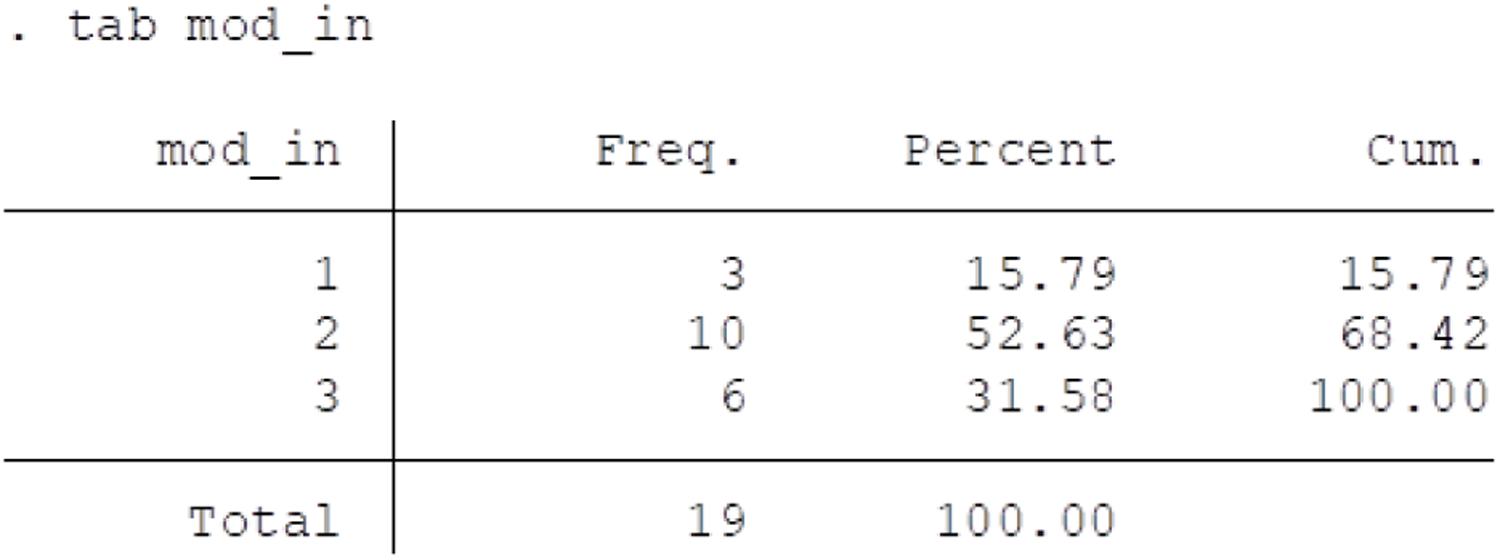

| mod_in | Freq. | Percent | Cum. |
| --- | --- | --- | --- |
| 1 | 3 | 15.79 | 15.79 |
| 2 | 10 | 52.63 | 68.42 |
| 3 | 6 | 31.58 | 100.00 |
| Total | 19 | 100.00 |  |

Vigorous PA intensity for 20 subjects

| vig_in | Freq. | Percent | Cum. |
| --- | --- | --- | --- |
| 1 | 13 | 72.22 | 72.22 |
| 2 | 4 | 22.22 | 94.44 |
| 3 | 1 | 5.56 | 100.00 |
| Total | 18 | 100.00 |  |

12 subjects

Low PA intensity for 12 subjects

| low_in | Freq. | Percent | Cum. |
| --- | --- | --- | --- |
| 1 | 2 | 18.18 | 18.18 |
| 2 | 1 | 9.09 | 27.27 |
| 3 | 8 | 72.73 | 100.00 |
| Total | 11 | 100.00 |  |

Moderate PA intensity for 12 subjects

| mod_in | Freq. | Percent | Cum. |
| --- | --- | --- | --- |
| 1 | 1 | 9.09 | 9.09 |
| 2 | 7 | 63.64 | 72.73 |
| 3 | 3 | 27.27 | 100.00 |
| Total | 11 | 100.00 |  |

Vigorous PA intensity for 12 subjects

| vig_in | Freq. | Percent | Cum. |
| --- | --- | --- | --- |
| 1 | 7 | 70.00 | 70.00 |
| 2 | 2 | 20.00 | 90.00 |
| 3 | 1 | 10.00 | 100.00 |
| Total | 10 | 100.00 |  |

8 subjects

Low PA intensity for 8 subjects

| low_in | Freq. | Percent | Cum. |
| --- | --- | --- | --- |
| 1 | 6 | 75.00 | 75.00 |
| 3 | 2 | 25.00 | 100.00 |
| Total | 8 | 100.00 |  |

Moderate PA intensity for 8 subjects

| mod_in | Freq. | Percent | Cum. |
| --- | --- | --- | --- |
| 1 | 2 | 25.00 | 25.00 |
| 2 | 3 | 37.50 | 62.50 |
| 3 | 3 | 37.50 | 100.00 |
| Total | 8 | 100.00 |  |

Vigorous PA intensity for 8 subjects

| vig_in | Freq. | Percent | Cum. |
| --- | --- | --- | --- |
| 1 | 6 | 75.00 | 75.00 |
| 2 | 2 | 25.00 | 100.00 |
| Total | 8 | 100.00 |  |

Fisher’s test comparing intensity by subjects included/excluded in analysis Low: 0.04

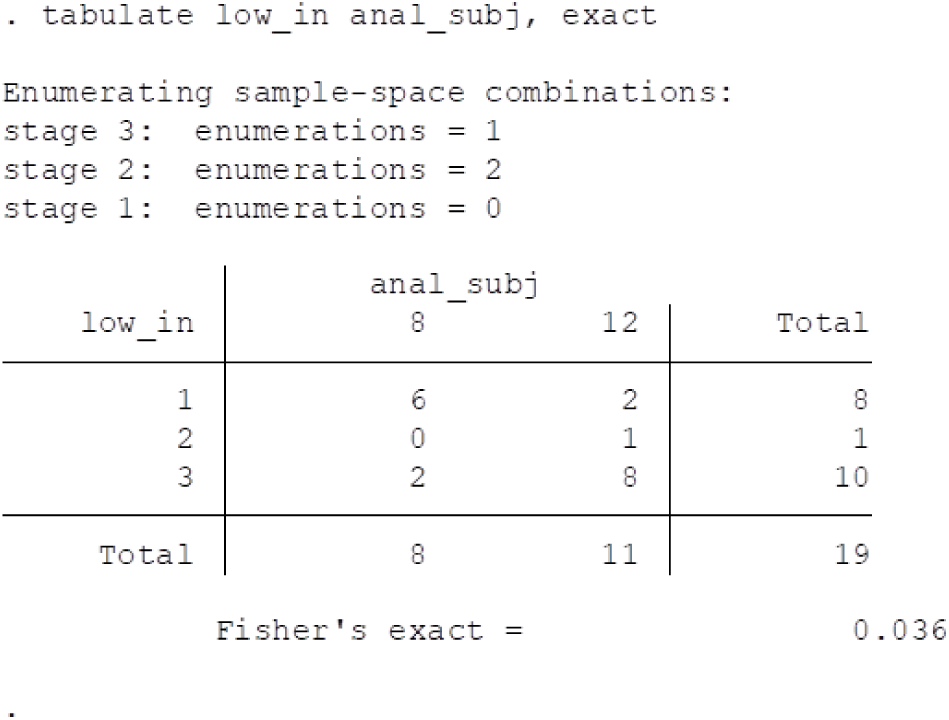

| low_in | anal_subj |  | Total |
| --- | --- | --- | --- |
|  | 8 | 12 |  |
| 1 | 6 | 2 | 8 |
| 2 | 0 | 1 | 1 |
| 3 | 2 | 8 | 10 |
| Total | 8 | 11 | 19 |

Fisher's exact = 0.036
``` Enumerating sample-space combinations: stage 3: enumerations = 1 stage 2: enumerations = 2 stage 1: enumerations = 0 ```

Moderate: 0.56

| mod_in | anal_subj |  | Total |
| --- | --- | --- | --- |
|  | 8 | 12 |  |
| 1 | 2 | 1 | 3 |
| 2 | 3 | 7 | 10 |
| 3 | 3 | 3 | 6 |
| Total | 8 | 11 | 19 |

Fisher's exact = 0.558

Vigorous: 1.0

| vig_in | anal_subj |  | Total |
| --- | --- | --- | --- |
|  | 8 | 12 |  |
| 1 | 6 | 7 | 13 |
| 2 | 2 | 2 | 4 |
| 3 | 0 | 1 | 1 |
| Total | 8 | 10 | 18 |

Fisher's exact = 1.000

Exit survey (n=12)

Satisfaction with HH walking recommendations

| Variable | Obs | Mean | Std. Dev. | Min | Max |
| --- | --- | --- | --- | --- | --- |
| ux5 | 11 | 3.545455 | .9341987 | 2 | 5 |

Ability to meet the physical activity needs: 4.27 (0.47)

| Variable | Obs | Mean | Std. Dev. | Min | Max |
| --- | --- | --- | --- | --- | --- |
| ux13 | 11 | 4.272727 | .4670994 | 4 | 5 |

Enjoy HH watch: 3.36 (1.43)

| Variable | Obs | Mean | Std. Dev. | Min | Max |
| --- | --- | --- | --- | --- | --- |
| ux34_watch | 11 | 3.363636 | 1.433369 | 0 | 5 |

Exit survey (n=8)

Walking: 4.4 (0.89)

| Variable | Obs | Mean | Std. Dev. | Min | Max |
| --- | --- | --- | --- | --- | --- |
| ux6 | 5 | 4.4 | .8944272 | 3 | 5 |

Ability to meet the physical activity needs: 4.2 (0.84)

| Variable | Obs | Mean | Std. Dev. | Min | Max |
| --- | --- | --- | --- | --- | --- |
| ux13 | 5 | 4.2 | .83666 | 3 | 5 |

| Age_new | anal_subj |  | Total |
| --- | --- | --- | --- |
|  | 8 | 12 |  |
| 18 | 0 | 2 | 2 |
| 19 | 1 | 1 | 2 |
| 20 | 1 | 2 | 3 |
| 21 | 1 | 1 | 2 |
| 22 | 2 | 2 | 4 |
| 23 | 0 | 1 | 1 |
| 25 | 1 | 0 | 1 |
| 27 | 0 | 1 | 1 |
| 33 | 1 | 0 | 1 |
| 34 | 1 | 0 | 1 |
| 35 | 0 | 1 | 1 |
| 37 | 0 | 1 | 1 |
| Total | 8 | 12 | 20 |

Fisher's exact = 0.919
Enumerating sample-space combinations:

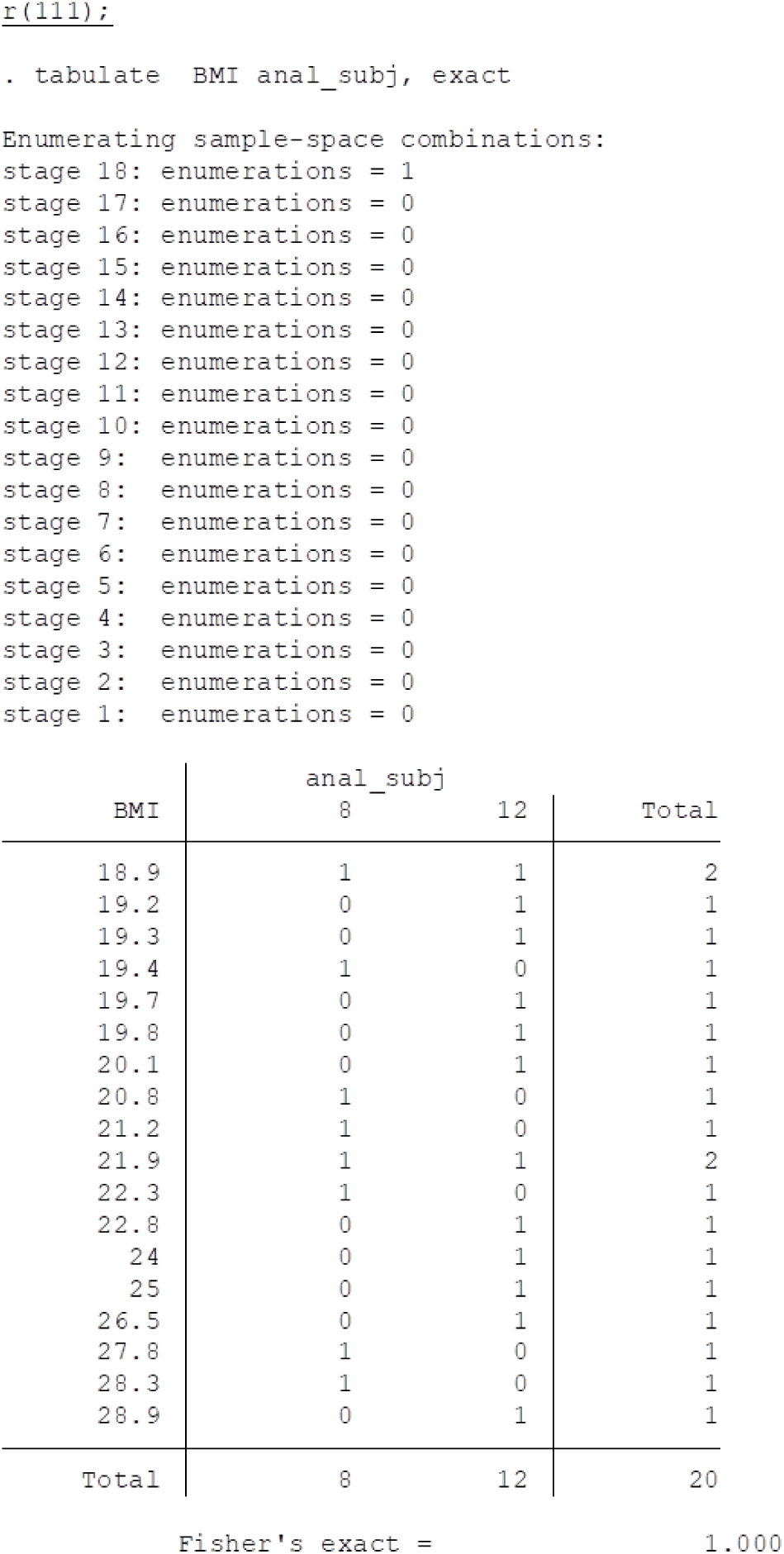

| BMI | anal_subj |  | Total |
| --- | --- | --- | --- |
|  | 8 | 12 |  |
| 18.9 | 1 | 1 | 2 |
| 19.2 | 0 | 1 | 1 |
| 19.3 | 0 | 1 | 1 |
| 19.4 | 1 | 0 | 1 |
| 19.7 | 0 | 1 | 1 |
| 19.8 | 0 | 1 | 1 |
| 20.1 | 0 | 1 | 1 |
| 20.8 | 1 | 0 | 1 |
| 21.2 | 1 | 0 | 1 |
| 21.9 | 1 | 1 | 2 |
| 22.3 | 1 | 0 | 1 |
| 22.8 | 0 | 1 | 1 |
| 24 | 0 | 1 | 1 |
| 25 | 0 | 1 | 1 |
| 26.5 | 0 | 1 | 1 |
| 27.8 | 1 | 0 | 1 |
| 28.3 | 1 | 0 | 1 |
| 28.9 | 0 | 1 | 1 |
| Total | 8 | 12 | 20 |

``` Fisher's exact = 1.000 ```

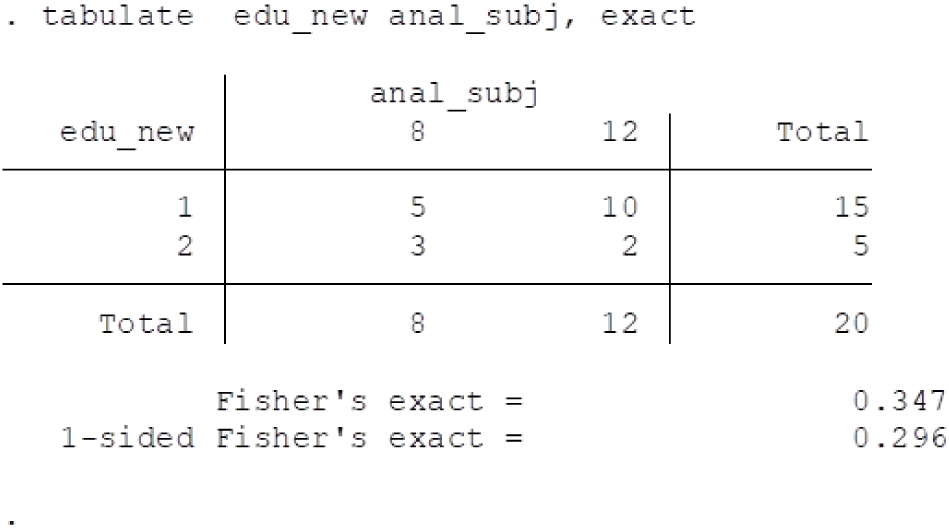

| edu_new | anal_subj |  | Total |
| --- | --- | --- | --- |
|  | 8 | 12 |  |
| 1 | 5 | 10 | 15 |
| 2 | 3 | 2 | 5 |
| Total | 8 | 12 | 20 |

| insurance_c | anal_subj |  | Total |
| --- | --- | --- | --- |
|  | 8 | 12 |  |
| 1 | 0 | 5 | 5 |
| 2 | 2 | 1 | 3 |
| 3 | 6 | 6 | 12 |
| Total | 8 | 12 | 20 |

Fisher's exact = 0.125
Enumerating sample-space combinations:
stage 3: enumerations = 1

| MaritalPartnershipSta<br>tus | anal_subj |  | Total |
| --- | --- | --- | --- |
|  | 8 | 12 |  |
| Married or Domestic.. | 0 | 1 | 1 |
| Significant Other | 3 | 5 | 8 |
| Single | 5 | 6 | 11 |
| Total | 8 | 12 | 20 |

Fisher's exact = 1.000

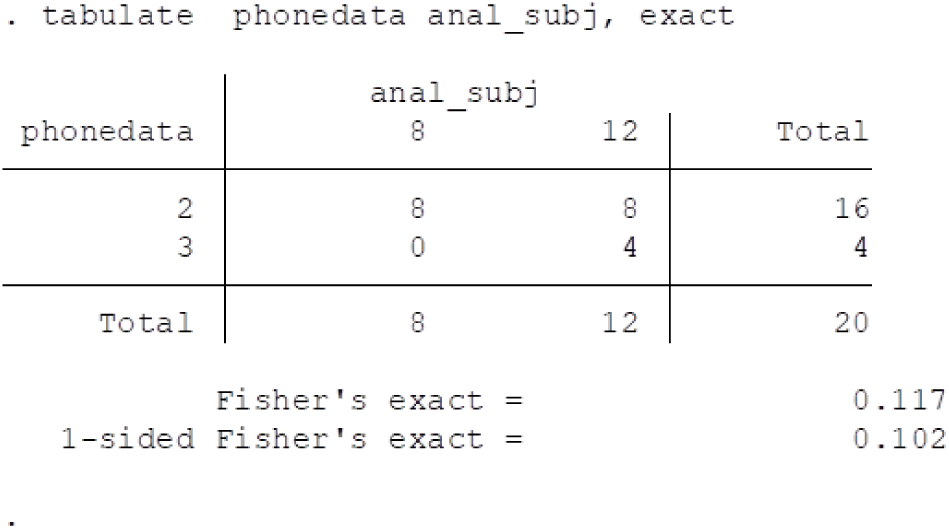

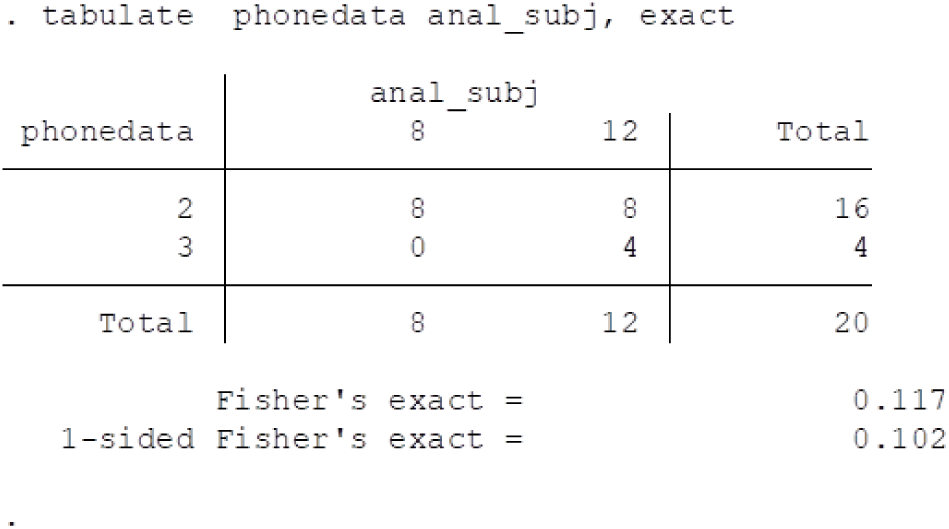

| wifi | anal_subj |  | Total |
| --- | --- | --- | --- |
|  | 8 | 12 |  |
| 1 | 8 | 12 | 20 |
| Total | 8 | 12 | 20 |

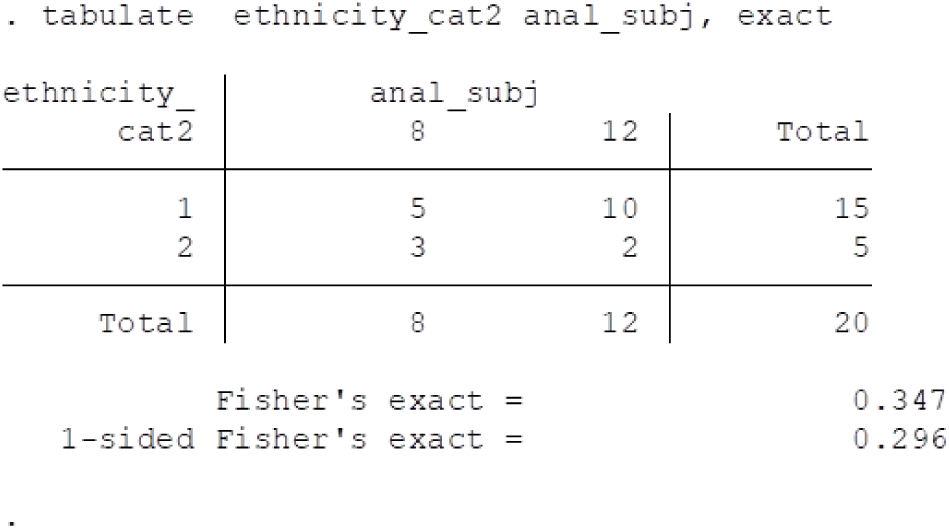

## Notes

### Competing Interest Statement

The authors have declared no competing interest.

### Author Declarations

University of California, Irvine, The IRB number: 2019-5363

